# Nationally representative prevalence and determinants of post-acute sequelae of SARS-CoV-2 infection (Long COVID) amongst Mexican adults in 2022

**DOI:** 10.1101/2023.07.10.23292475

**Authors:** Omar Yaxmehen Bello-Chavolla, Carlos A. Fermín-Martínez, Luisa Fernández-Chirino, Daniel Ramírez-García, Arsenio Vargas-Vázquez, Martín Roberto Basile-Alvarez, Paulina Sánchez Castro, Alejandra Núñez-Luna, Neftali Eduardo Antonio-Villa

## Abstract

**OBJECTIVE:** To characterize the epidemiology of post-acute sequelae after SARS-CoV-2 infection (PASC) in Mexico during 2022 and identify potential predictors of PASC prevalence using nationally representative data.

**METHODS:** We analyzed data from the 2022 Mexican National Health and Nutrition Survey (ENSANUT) totaling 24,434 participants, representing 85,521,661 adults ≥20 years. PASC was defined using both the World Health Organization definition and a PASC score ≥12. Estimates of PASC prevalence were stratified by age, sex, rural vs. urban setting, social lag quartiles, number of reinfections, vaccination status and by periods of predominance of SARS-CoV-2 circulating variants. Predictors of PASC were assessed using logistic regression models adjusted by survey weights.

**RESULTS:** Persistent symptoms after SARS-CoV-2 infection were reported by 12.44% (95%CI 11.89-12.99) of adults ≥20 years in Mexico during 2022. The most common persistent symptoms were musculoskeletal pain, headache, cough, loss of smell or taste, fever, post-exertional malaise, brain fog, anxiety, chest pain, and sleep disorders. PASC was present in 21.21% (95%CI 7.71-9.65) subjects with previously diagnosed COVID-19. Over 28.6% patients with PASC reported symptoms persistence ≥6 months and 14.05% reported incapacitating symptoms. Higher PASC prevalence was associated with SARS-CoV-2 reinfections, depressive symptoms and living in states with high social lag. PASC prevalence, particularly its more severe forms, decreased with COVID-19 vaccination and for infections during periods of Omicron variant predominance.

**CONCLUSIONS:** PASC implies a significant public health burden in Mexico as the COVID-19 pandemic transitions into endemicity. Promoting reinfection prevention and booster vaccination may be useful to reduce PASC burden.

## INTRODUCTION

Post-acute persistent symptoms of severe acute coronavirus 2 (SARS-CoV-2) infection remain a challenge to reduce the burden of SARS-CoV-2 in affected patients as the coronavirus disease (COVID-19) pandemic progresses towards endemicity (1). The definition of post-acute sequelae after SARS-CoV-2 infection (PASC) is still evolving given heterogeneous patient profiles, with other terms also recognizing it as post-acute COVID-19 condition, long COVID, and post-acute COVID-19 syndrome (2). A Delphi consensus by the World Health Organization defined it as a condition affecting individuals with suspected or confirmed SARS-CoV-2 infection exhibiting persistent symptoms lasting for at least two months without an alternative pathophysiological explanation (3). Recently, Thaweethai et al. developed a self-reported symptom-based definition to better characterize persistent COVID-19 symptoms across cohorts and provide another overview of the epidemiology of PASC (4). To this date, reports on the epidemiology of persistent symptoms are scarce, particularly in Latin American countries, which hinders recognition of the magnitude of the problem and implementation of public policies aimed at reducing PASC burden (5).

Mexico is one of the countries with the highest morbidity and mortality related to COVID-19 world-wide (6). A landscape of high cardio-metabolic burden with marked social inequalities increased the impact of the pandemic and led to sustained excess deaths, which were ameliorated with the availability of vaccines (7–9). Despite the well-characterized acute impact of the SARS-CoV-2 pandemic in low- and middle-income settings, information regarding the impact of debilitating chronic sequelae of SARS-CoV-2 infection in these settings is lacking. In Mexico, few reports have evaluated the impact of PASC, particularly focusing on cases with severe COVID-19 (10,11). However, nationally representative estimates of persistent COVID-19 symptoms and particularly PASC as defined by current criteria in Mexico remain unreported. Here, we aimed to estimate the prevalence of persistent COVID-19 symptoms and post-acute sequelae of SARS-CoV-2 infection (PASC) using a nationally representative sample of Mexican adults for the year 2022. We also aimed to evaluate potential correlates of PASC prevalence including sociodemographic and infection-related determinants to characterize vulnerable groups and potential areas of intervention to ameliorate the long-term burden of COVID-19 in Mexico.

## METHODS

### Study design

We analyzed data from the Mexican National Health and Nutrition Survey (ENSANUT) for the year 2022. Briefly, ENSANUT is a population-based survey that aims to assess the health and nutritional status of Mexican adults. ENSANUT is representative at a national, regional, and rural/urban level, as it uses two-stage probabilistic cluster stratified sampling based on households and individuals. Participants underwent a comprehensive questionnaire collecting demographic, socioeconomic, and health-related data, and a physical exam including measurement of blood pressure and anthropometry. ENSANUT 2022 was collected from July to December 2022 in 10,160 households (response rate of 73%) which amounts to 11,472 complete interviews in adults ≥20 years old (12). This survey additionally aimed to investigate health and well-being of Mexican adults during the COVID-19 pandemic and a random subsample of 5,971 adults ≥20 years old provided a capillary blood sample to detect antibodies for SARS-CoV-2 infection. A second subsample (n=2,092) had an additional biochemical evaluation with venous blood samples for glycated hemoglobin (HbA1c), fasting glucose, and a fasting lipid profile. A complete flowchart of participant selection is outlined in **Supplementary Materials**.

### SARS-CoV-2 seroprevalence

Seropositivity to SARS-CoV-2 was evaluated using an Elecsys detection assay for IgG against the N-protein (SARS-CoV-2 Nucleocapsid, #09 203 079 190, Roche-N) validated by the Institute for Epidemiological and Diagnostic Reference. Samples were considered positive if quantification was ≥0.72.0 U/ml (13). COVID-19 seroprevalence was estimated using population sample weights with the *survey* R package.

### Variable definitions

#### Persistent COVID-19 symptoms

Previously diagnosed COVID-19 was defined by self-report among individuals who answered, “at least once” to the question: “Since February 2020, how many times have you been diagnosed with COVID-19 by health personnel?” Amongst individuals with at least one previous COVID-19 diagnosis, persistent symptoms after SARS-CoV-2 infection were asked (3) using the question: “Regarding the last time you had COVID, did you continue to present any of these symptoms / sequelae one month after your illness began?” and interrogated the presence of the following symptoms: cough, fatigue or tiredness, anxiety, depression, fever, difficulty sleeping, kidney complications, loss of appetite, weight loss, headache, dizziness, muscle or joint pain, difficulty breathing (dyspnea), shortness of breath, chest pain, vomiting or diarrhea (gastrointestinal symptoms), loss or decrease in smell, loss or decrease in taste, difficulty thinking or concentrating or other symptoms, including post-exertional malaise, which we defined as symptoms which worsen with activity and impede normal functioning. Participants were also asked to clarify if the symptoms persisted for less than an additional month, from one to three months, from three to six months, more than 6 months, or are still present.

#### Post-acute COVID-19 sequelae

We used the definition reached by Delphi consensus from the World Health Organization, which defines PASC, which defines it as the presence of at least one persistent symptom, in individuals with previously diagnosed SARS-CoV-2 infection, and which referred persistent symptoms lasting ≥3 months from the previous COVID-19 diagnosis. Based on this, we classified persistent symptoms into subacute or ongoing symptomatic COVID-19, defined as the presence of any persistent symptom 4-12 weeks beyond acute COVID-19 and PASC(14). Furthermore, based on a recent definition by Thaweethai et al. we calculated the PASC score, which defined the presence of PASC with a score ≥12 points to assess its prevalence in this population (4). Symptoms such as palpitations, chronic thirst, decreased sexual desire or capacity, or abnormal movements were not considered in ENSANUT 2022 and were not included for score calculation. This complementary definition was included as a sensitivity analysis and was considered irrespective of symptom duration.

#### Modifying factors

We evaluated age categories of 10-year intervals (20-29, 30-39, 40-49, 50-59, 60-69 or ≥70 years), sex, rural/urban area, smoking status and presence of diabetes and hypertension as modifying factors of PASC prevalence. The following factors were also considered to evaluate components of SARS-CoV-2 infection:

- **SARS-CoV-2 reinfection** – Defined as having been diagnosed with COVID-19 more than one time by healthcare personnel (self-report).
- **Predominant SARS-CoV-2 variants** – SARS-CoV-2 infection was assumed to be most likely caused by the predominant variant based on the date of symptom onset. Based on data submitted to GISAID (48), from March 3rd, 2020, until December 30th, 2020, the predominant SARS-CoV-2 variant was the ancestral strain, followed by the predominance of the B.1.1.519 variant until June 6th, 2021, the B.1.617.2 (Delta) variant until December 6th, 2021. B.1.1.529.1 (Omicron) subvariant was considered predominant from December 7th, 2022, onwards (15). For modeling, COVID-19 were classified into those likely caused by the Omicron variant or otherwise.
- **COVID-19 vaccination –** Vaccination was defined by self-report among individuals who answered “yes” to the question: “Have you been vaccinated for COVID-19?” and further questioned if receiving one, two, three or four vaccine doses.
- **Social lag –** We used data from the National Council for Evaluation of Social Development Policy (CONEVAL), which provides state-level estimates of the 2020 social lag index (SLI), which is a composite measure of access to education, health care, dwelling quality, and basic services in Mexico (16). To assess marginalization independently of urbanization, we extracted mean population density from each state and regressed it onto SLI and obtained the residuals which represent a density-independent social lag index (DISLI) (9,17,18). We then categorized states into DISLI quartiles (Q1-Q4) and classified individuals from each quartile to each according to their state of residence.
- **Depressive symptoms –** Evaluated using he seven-item version of the Center for Epidemiologic Studies Depression Scale (CESD-7). CESD-7 measures the frequency with which depressive symptoms are experienced during the week prior to the interview collected at ENSANUT 2022. Cut-offs to identify the presence of moderate or severe depressive symptoms were ≥9 points for adults aged 20-59, and ≥5 points for adults ≥60 years (19,20).
- **Incapacitating symptoms –** Defined if the person answered “yes” to the question “Do these [persistent COVID-19] symptoms prevent you or did they prevent you from taking care of yourself? For example, that difficulties to bathe or dress oneself”.

### Statistical analyses

#### Weighted prevalence of PASC and persistent COVID-19 symptoms

Prevalence of PASC and persistent symptoms were estimated using sample weights from ENSANUT for participants ≥20 years, as well as those with SARS-CoV-2 seropositivity and for the population with previously diagnosed COVID-19; all estimations were conducted using the *survey* R package (21). We further performed weighted subgroup estimation for prevalence trends stratified by age category, sex, geographical region, previous reinfection, vaccination status, infection during periods of Omicron variant predominance, indigenous identity, rural or urban area, and DISLI category (high or low/middle).

#### Predictors of PASC prevalence

To identify correlates of PASC positivity we fitted fixed effects logistic regression models considering survey weights amongst individuals with previously diagnosed COVID-19 and complete data including comorbidities. A sensitivity analysis was conducted exploring predictors for any sequelae present compared to the PASC definition. All statistical analyses were conducted using R version 4.1.2 and p-values thresholds are estimated for a two-sided significance level of α = 0.05.

## RESULTS

### Study population

We included adults aged ≥20 years old who completed the health questionnaire, totaling 24,434 participants (expanded to 85,521,661 adults); amongst them, 4,898 participants were selected for capilar blood sampling to estimate SARS-CoV-2 seropositivity (expanded to 85,098,924 adults). A flowchart diagram detailing the participant selection process is available in **Supplementary Figure 1**. Overall, we identified a prevalence of previously diagnosed COVID-19 amongst adults ≥20 years of 22.0% (95%CI 21.31-22.69), which contrasted markedly with the seroprevalence of IgG against the N-protein of SARS-CoV-2, estimated at 93.66% (95%CI 92.78-94.54). Overall, approximately 86.21% (95%CI 85.64-86.78) of adults had received at least one dose of COVID-19 vaccines, with 11.62% (95%CI 11.1-12.14) receiving only one dose, 29.6% (95%CI 28.83-30.37) with two doses, 39.51% (95%CI 38.73-40.29) with three doses and 5.48% (95%CI 5.11-5.85) with four doses.

### Prevalence of persistent COVID-19 symptoms

Overall, we identified that 12.44% (95%CI 11.89-12.99) of adults ≥20 years had at least one persistent COVID-19 symptom for ≥1 month, which equates to a total of 10,634,663 individuals (95%CI 10,155,424-11,113,902). Amongst them, 45.87% (95%CI 43.49-48.25) lasted <1 additional month, 16.74% (95%CI 14.93-18.55) persisted between 1-3 months, 4.54% (95%CI 3.56 -5.52) between 3-6 months, 4.26% (95%CI 3.28-5.24) for >6 months and 28.6% (95%CI 26.43-30.77) persisting until the date of the interview. The prevalence of any persistent symptoms was similar when analyzing only N-protein seropositive adults (12.7%, 95%CI 11.48-14.06) and was higher when considering only cases with previously diagnosed COVID-19 by a physician (56.52%, 95%CI 54.75-58.29). The ten most frequent persistent symptoms amongst subjects with SARS-CoV-2 N-protein seropositivity were musculoskeletal pain, headache, cough, loss of smell or taste, fever, post-exertional malaise, brain fog, anxiety, chest pain, and sleep disorders (**Supplementary Material**).

### Persistent symptoms in selected subgroups

Amongst SARS-CoV-2 N-protein seropositive individuals subacute or ongoing symptomatic COVID-19 was present in 7.42% (95%CI 6.42-8.42), whilst PASC was present in 5.51% (95%CI 4.61-6.41) of cases (**Figure 1)**. The five most common persistent symptoms in subacute COVID-19 were cough, headache, fever, musculoskeletal pain, and loss of smell or taste, whilst for PASC they were musculoskeletal pain, post-exertional malaise, headache, cough, and dyspnea (**Figure 1A-B**). Regarding sex, the prevalence of any persistent symptom amongst SARS-CoV-2 N-protein seropositive individuals was higher for males (14.12%, 95%CI 12.42-15.85) compared to females (10.50%, 95%CI 8.56-12.44); in males the five most frequent persistent symptoms were headache, musculoskeletal pain, and cough, whilst for females they were cough, musculoskeletal pain, and fever (**Supplementary Material**). In cases with previously diagnosed COVID-19 SARS-CoV-2 primoinfection was associated with lower prevalence of persistent sequelae (55.39%, 95%CI 53.49-57.29) compared to cases with at least one SARS-CoV-2 reinfection (64.41%, 95%CI 59.69-69.13); in cases with SARS-CoV-2 primoinfection the most common persistent symptoms were musculoskeletal pain, headache, cough, post-exertional malaise, and loss of smell or taste, whilst for reinfection they were cough, headache, post-exertional malaise, musculoskeletal pain, and fever (**Supplementary Material**).

**Figure 1.**
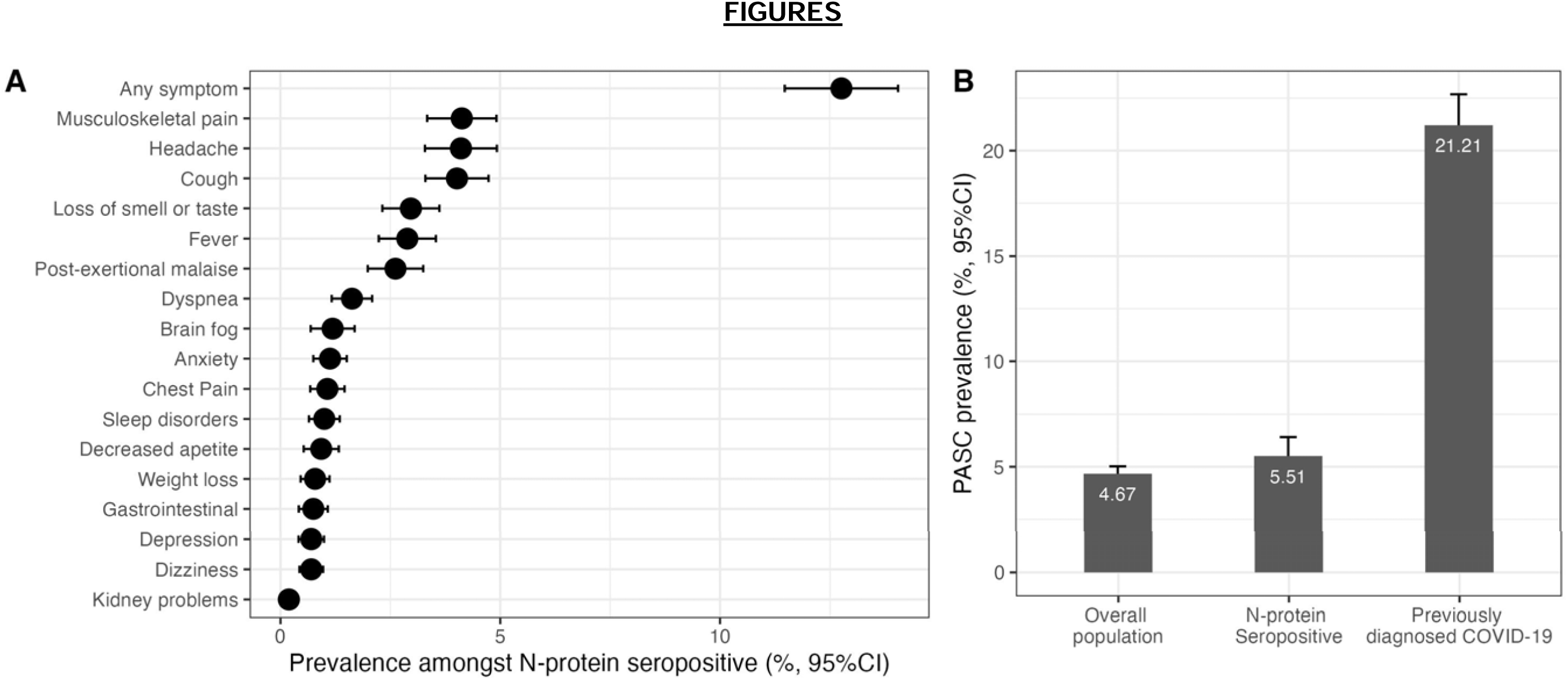
Prevalence of persistent COVID-19 symptoms amongst SARS-CoV-2 N-protein seropositive adults ≥20 years in ENSANUT 2022 (**A)** and prevalence of post-acute sequelae of SARS-CoV-2 symptoms (PASC) as identified by a PASC score ≥12 in the overall population, in SARS-CoV-2 N-protein seropositive adults and in individuals with COVID-19 previously diagnosed by a physician in the ENSANUT 2022 sample (**B**).

### Prevalence of PASC in Mexico

Using the WHO definition, we identified a prevalence of 4.67% (95%CI 4.32-5.02) in the overall population, 5.51% (95%CI 4.61-6.41) amongst SARS-CoV-2 N-protein seropositive population, and 21.21% (95%CI 19.74-22.68) in subjects with previously diagnosed COVID-19, totaling 3,990,011 affected adults (95%CI 3,685,878-4,294,144; **Supplementary Material**). We identified the highest prevalence of PASC amongst subjects with previously diagnosed COVID-19 in Mexico City/Mexico State and the Peninsula region, whilst the lowest prevalence was observed in the US Border region, coincidentally most clustering in Northern Mexico (**Figure 2A**). When comparing characteristics between individuals with and without PASC, we identified that individuals with PASC clustered most persistent symptoms, even those not included in the PASC score; furthermore, higher rates of reinfections, diabetes and hypertension were observed individuals with PASC (**Table 1**). Overall, 15.13% (95%CI 13.50-16.76) of individuals affected by persistent symptoms reported incapacity to perform everyday tasks including take care of oneself; this rate was 14.06% (95%CI 11.37-16.75) amongst PASC compared to 7.13% (95%CI 6.17-8.09) in individuals without PASC who had previously diagnosed with COVID-19, indicating a high burden of PASC on everyday functioning. Finally, we analyzed cases with PASC score ≥12, observing a prevalence of 1.91% (95%CI 1.69-2.13) in the general population, 2.02% (95%CI 1.48-2.56) in SARS-CoV-2 N-protein seropositive individuals and 8.68% (95%CI 7.72-9.65) in individuals with previously diagnosed SARS-CoV-2 infection. Notably, incapacitating PASC symptoms were observed in 45.38% (95%CI 39.65-51.11) of subjects with PASC score ≥12, indicating a more severe phenotype. Furthermore, comparing PASC positive (score ≥12) vs. PASC indeterminate subjects (score <12) reinfection rates and symptom clustering occurred most frequently in PASC positive subjects (**Supplementary Material)**; interestingly, lower vaccination and infections detected during periods of Omicron variant predominance were observed in PASC positive vs. PASC indeterminate individuals. This indicated that the PASC score likely detected individuals with a more severe PASC phenotype.

**Figure 2.**
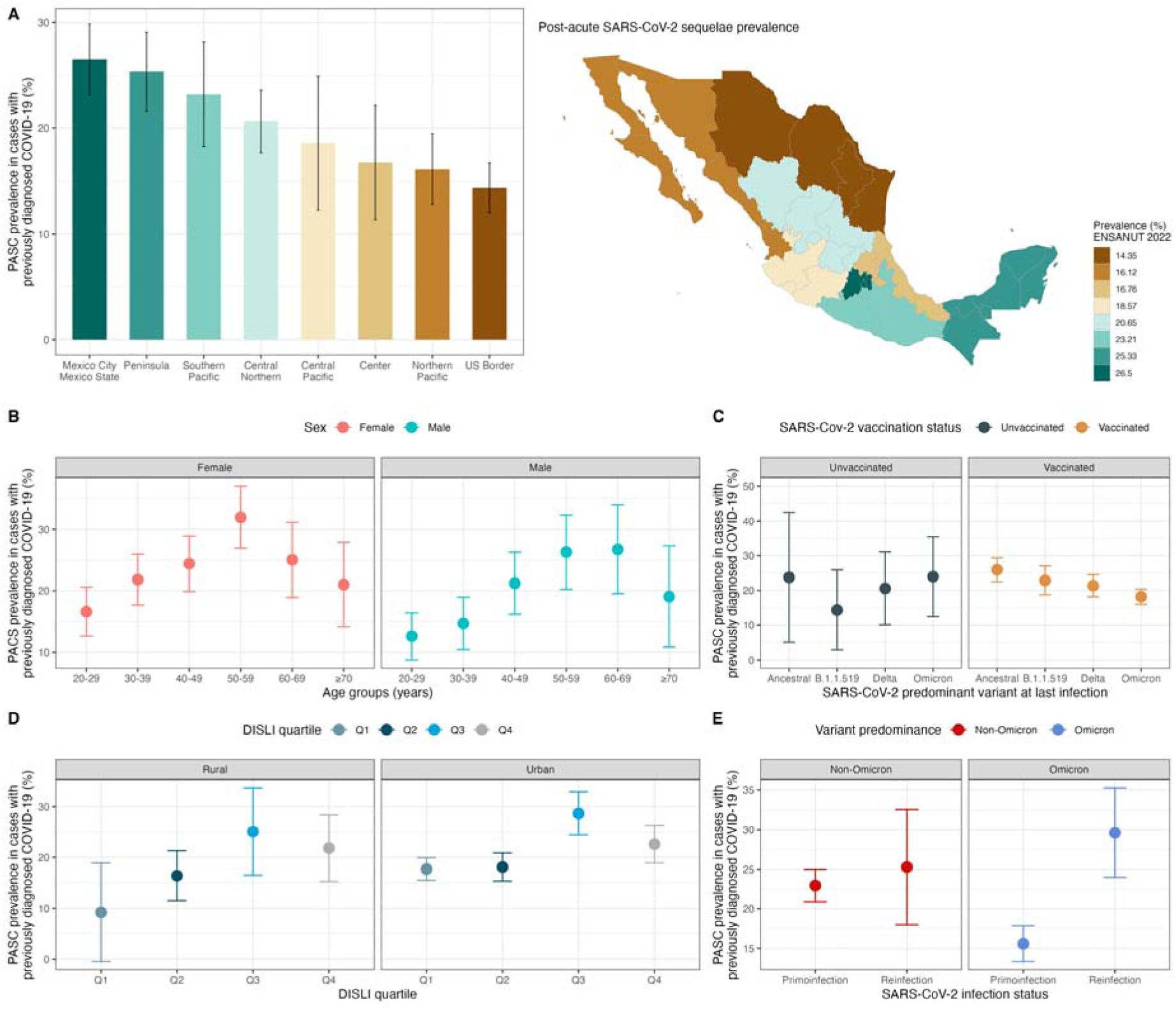
Prevalence of post-acute sequelae of SARS-CoV-2 symptoms (PASC) as identified by a PASC score ≥12 stratified by geographical regions in Mexico within the ENSANUT 2022 sample of adults ≥20 years (**A**). The figure also shows PASC prevalence stratified by age and sex categories (**B**), by predominant circulating variant at the time of last SARS-CoV-2 infection and vaccination status (**C**), by density-independent social lag index (DISLI) quartiles in rural vs. urban setting (D), and in subjects with primoinfection compared to at least one SARS-CoV-2 reinfection during periods of non-Omicron and Omicron variant predominance (**E**).

**TABLE 1.**
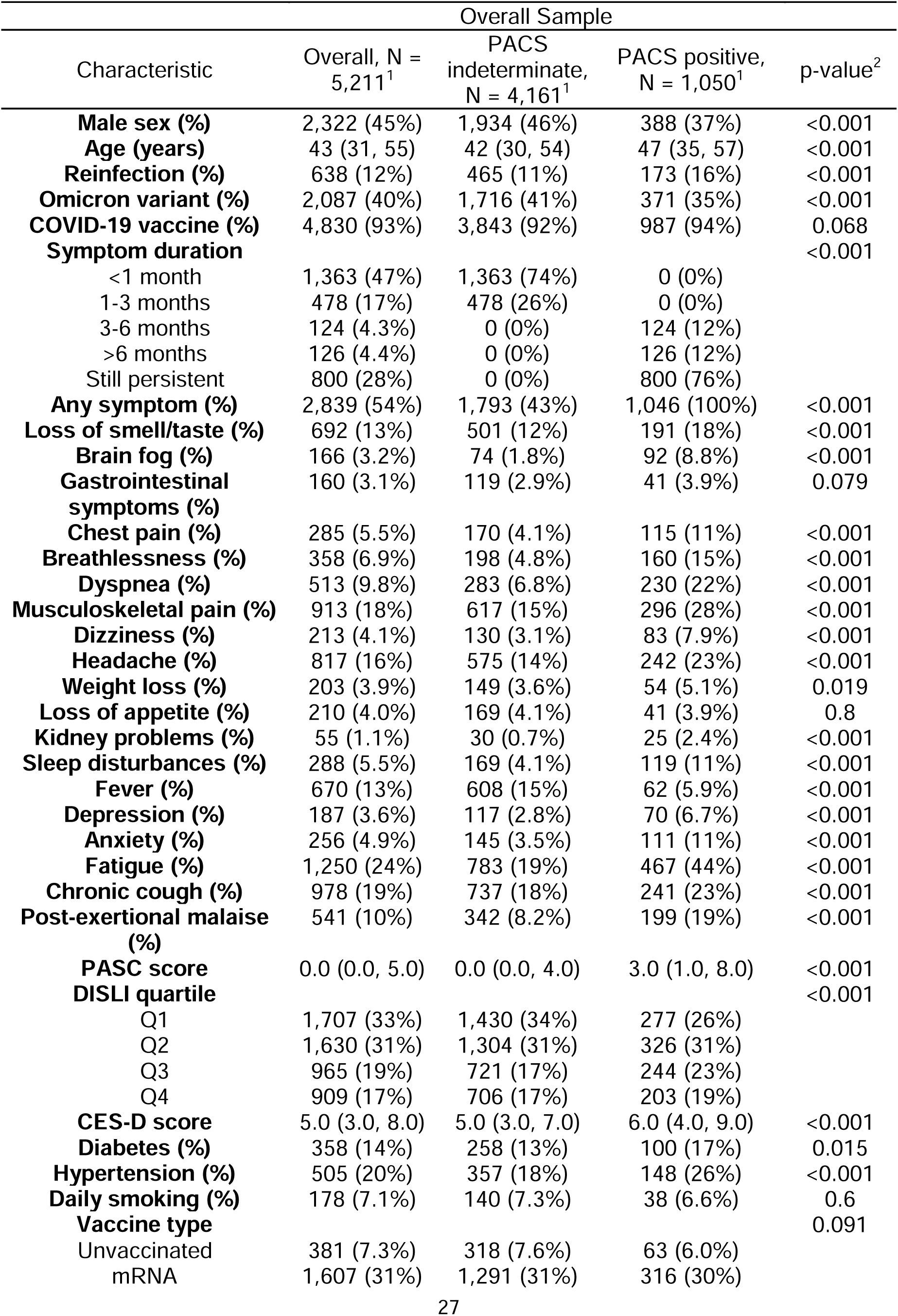
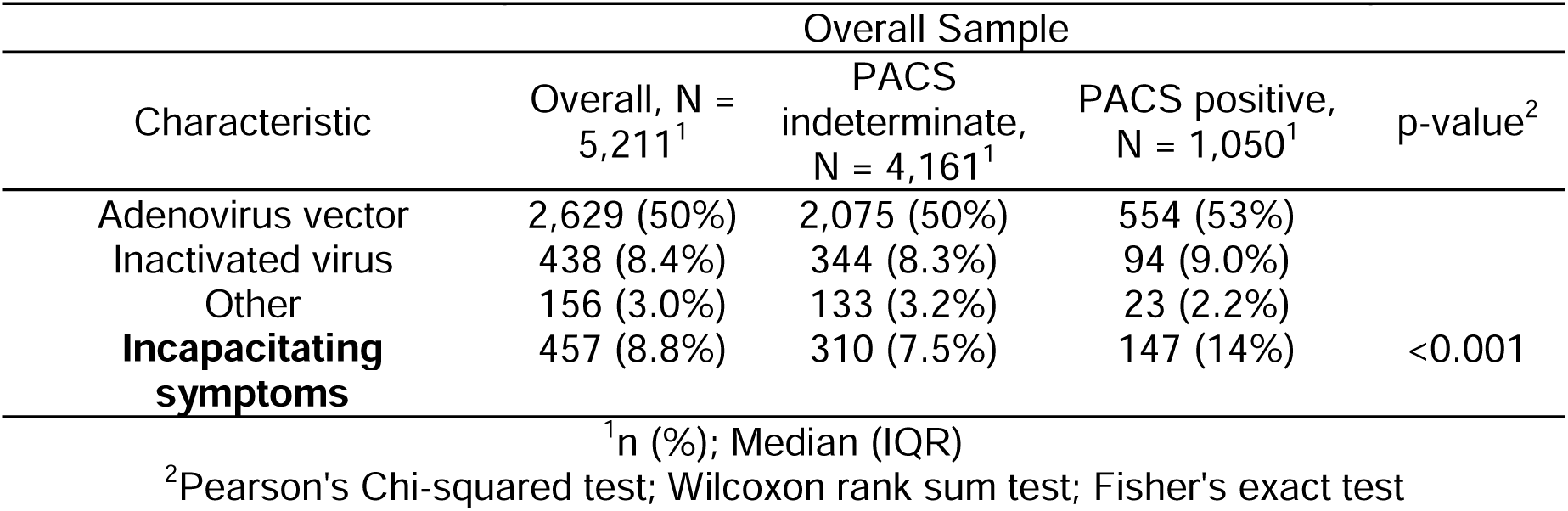
Sociodemographic and clinical characteristics of individuals with post-acute sequelae of SARS-CoV-2 symptoms (PASC) as defined by the World Health Organization compared individuals without PASC and previously diagnosed with COVID-19 in the ENSANUT 2022 sample.

### PASC prevalence in selected subgroups

PASC prevalence estimates generally increased with age and were higher in females compared to males (**Figure 2B**); furthermore, PASC prevalence was higher with increasing DISLI values, particularly in urban settings (**Figure 2D**). For infection-related variables the prevalence of PASC was lower if the last infection occurred during periods of high Omicron variant circulation compared to other SARS-CoV-2 variants and was generally lower in vaccinated subjects irrespective of variant predominance (**Figure 2C)**; similarly, prevalence of PASC was higher in subjects with at least one reinfection compared to primoinfection, but this was more evident in cases where COVID-19 was acquired during periods of Omicron variant predominance (**Figure 2E**). Finally, when evaluating PASC prevalence per year at which the last SARS-CoV-2 infection was diagnosed, prevalence changed from 25.82% (95%CI 22.40-29.24) for cases infected in 2020, to 21.96% for cases infected in 2021 (95%CI 19.65-24.27), and to 17.60% (95%CI 15.36-19.84) for cases infected in 2022 (**Supplementary Material**).

### Modifiers of PASC prevalence

We fitted fixed effects logistic regression model considering survey weights to identify correlates of PASC positivity amongst individuals with previously diagnosed COVID-19 and complete data including comorbidities (n=2,504). We identified that infection during periods of Omicron variant predominance (OR 0.73, 95%CI 0.59-0.90) and male sex (OR 0.75, 95%CI 0.61-0.91) were associated with lower odds of PASC positivity. In addition, having had at least one SARS-CoV-2 reinfection (OR 1.83, 95%CI 1.38-2.41), having moderate to severe depressive symptoms as identified by CES-D (OR 1.89, 95%CI 1.51-2.36), older age, and living in states with higher social lag (OR 1.48, 95%CI 1.10-1.99 for Q4 vs Q1) were associated with higher odds of PASC positivity (**Figure 3A**). When evaluating the same predictors but using the definition of PASC score ≥12 points, which likely indicates a more severe PASC phenotype, infection during periods of Omicron variant predominance, COVID-19 vaccination, having had at least one SARS-CoV-2 reinfection, living in states with higher social lag, and moderate to severe depressive symptoms by CES-D were associated with the outcome (**Figure 3B)**; further disaggregating by the number of vaccine doses compared to unvaccinated individuals showed a similar protective effect with increasing doses and irrespective of vaccine platform for primary vaccination schedule (**Supplementary Material**).

**FIGURE 3.**
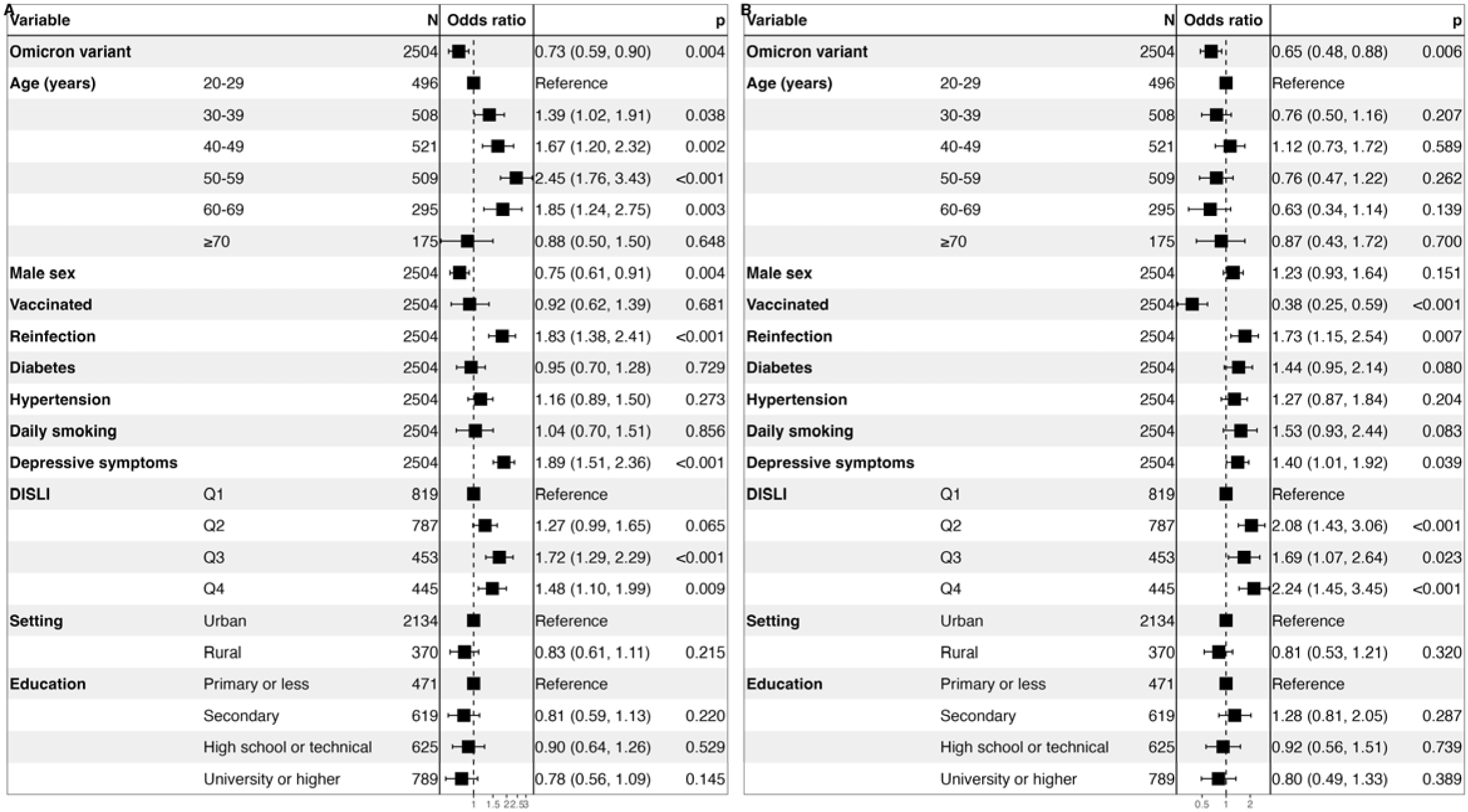
Fixed effects logistic regression model adjusted by survey weights for prediction of post-acute sequelae of SARS-CoV-2 symptoms (PASC) identified by a PASC score ≥12 (**A**) and the presence of any persistent COVID-19 symptom (**B**) amongst Mexican adults enrolled in ENSANUT 2022. <u>Abbreviations:</u> DISLI, Density-independent social lag-index; Depressive symptoms: Moderate to severe depressive symptoms identified by the Center for Epidemiologic Studies Depression Scale.

### Predictors of incapacitating PASC symptoms

Amongst individuals with persistent symptoms a PASC score ≥12 points resulted in higher rates of incapacitating symptoms at all time points and independent of the WHO PASC definition (**Figure 4A**). Predictors of incapacitating symptoms included previously diagnosed diabetes (OR 1.98, 95%CI 1.00-3.90), moderate to severe depressive symptoms (OR 1.85, 95%CI 1.07-3.20), age 50-59 years compared to younger individuals (OR 3.39, 95%CI 1.32-10.37), having had at least one SARS-CoV-2 reinfection (OR 2.58, 95%CI 1.24-5.31) and a lower risk observed in individuals infected during periods of Omicron variant predominance (OR 0.42, 95%CI 0.22-0.79). Notably, the most significant predictor of incapacitating PASC symptoms was a PASC score ≥12 points (OR 4.92, 95%CI 2.37, 10.12).

**FIGURE 4.**
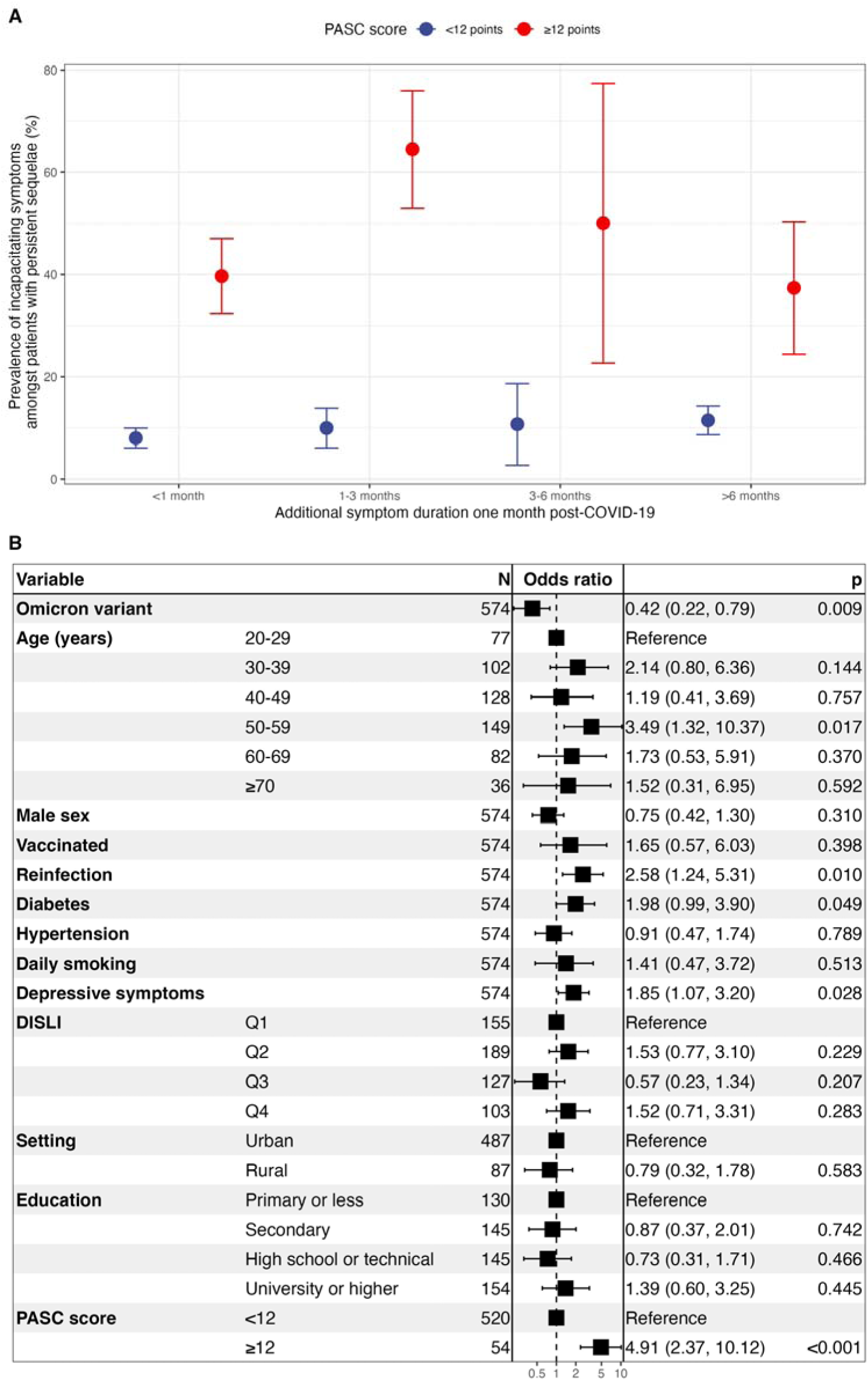
Prevalence of incapacitating symptoms amongst patients with persistent post-COVID-19 sequelae disaggregated by PASC scores using a 12-point threshold (**A**). The figure also shows a fixed effects logistic regression model adjusted by survey weights for prediction of debilitating PASC symptoms (**A**) and the presence of any persistent COVID-19 symptom (**B**) amongst Mexican adults enrolled in ENSANUT 2022. <u>Abbreviations:</u> PASC, Post-acute sequelae of SARS-CoV-2 infection; DISLI, Density-independent social lag-index; Depressive symptoms: Moderate to severe depressive symptoms identified by the Center for Epidemiologic Studies Depression Scale.

## DISCUSSION

Here, we conducted the first comprehensive report on the prevalence of PASC in Mexican adults during the year 2022, surveying a total of 24,434 participants. Persistent COVID-19 symptoms were reported by 12.44% of adults ≥20 years in Mexico and amongst 12.7% of adults with SARS-CoV-2 N-protein seropositivity during 2022. The most commonly reported persistent COVID-19 symptoms amongst SARS-CoV-2 N-protein seropositive adults include musculoskeletal pain, followed by headache, cough, loss of smell or taste, fever, post-exertional malaise, brain fog, anxiety, chest pain, and sleep disorders. Using a recent the WHO PASC definition (3) we identified a prevalence of 5.51% amongst SARS-CoV-2 N-protein seropositive Mexican adults and 21.21% amongst cases with previously diagnosed COVID-19; the prevalence of PASC increased with age and was higher for females, for SARS-CoV-2 infections detected in 2020 and in settings with higher social lag in urban settings. PASC was associated with increasing number of SARS-CoV-2 reinfections, moderate to severe depressive symptoms and living in states burdened by sociodemographic inequalities. We also confirm previous reports that COVID-19 vaccination and infection during periods of Omicron variant predominance decreases odds of severe forms of PASC (2,22,23). Notably, over 28.6% patients with PASC reported persistence of symptoms over 6 months and over 14.06% reported incapacitating symptoms, which indicate a subset of participants in whom interventions may lead to a significant improvement in quality of life (24). Finally, we identified that incapacitating PASC symptoms were associated with diabetes, moderate to severe depressive symptoms, SARS-CoV-2 reinfections and a PASC score ≥12 points. Our study represents the first comprehensive and nationally representative report of PASC in Mexican adults and the first report on the impact of vaccination, SARS-CoV-2 variants, reinfections, and sociodemographic inequalities in the prevalence of PASC and symptom persistence and severity in Mexico.

A previous systematic review identified that approximately 72.5% of individuals previously infected by SARS-CoV-2 experienced post-acute sequelae (25). This contrasts with a lower reported prevalence of 56.52% amongst individuals with previously diagnosed COVID-19 reported in our study and with the even lower estimate amongst SARS-CoV-2 seropositive adults. A previous study using data from ENSANUT 2020 reported that amongst SARS-CoV-2 seropositive individuals who recovered from COVID-19 in Mexico, approximately 15.7% reported unspecified sequelae, with these being higher in females, older adults and hospitalized individuals (11). In our study, we identified that the main predictors of higher PASC prevalence include SARS-CoV-2 reinfections, moderate to severe depressive symptoms and sociodemographic inequalities as proxied by the DISLI. Similar to our findings, previous reports from the UK showed that individuals with PASC clustered in areas of higher socioeconomic deprivation (26), supporting the notion that structural determinants of health may influence the incidence and natural history of chronic debilitating diseases including chronic fatigue syndrome, myalgic encephalitis and PASC (27,28), as it does during the acute phase of SARS-CoV-2 infection (9). Our findings are also in agreement with previous data supporting that SARS-CoV-2 reinfections increased the risk of SARS-CoV-2 sequelae, irrespective of predominant variant; although, lower risk of PASC has been consistently associated with infections caused by the Omicron compared to previous circulating variants (29,30). Finally, the higher rate of moderate to severe depressive symptoms and self-reported anxiety in PASC indicates a relevant area of intervention for affected individuals, particularly since long-term neuropsychiatric sequelae has been consistently reported in SARS-CoV-2 convalescent individuals after recovery from acute infection (4,14,31–33).

The precise nature behind the pathophysiology of PASC is unclear. Current evidence suggests involvement of SARS-CoV-2 mediated pathophysiological changes in several tissues, immune system dysregulation and immune response viral particle persistence and expected sequelae of critical illness. Increased inflammatory responses and an inadequate adaptation of the hypothalamic–pituitary–adrenal axis to hypocortisolemia after acute SARS-CoV-2 infection may underlie the occurrence of symptoms related to brain fog, post-exertional malaise, and neuropsychiatric symptoms, including anxiety and depression in PASC positive patients (2,34–36), which often last the longest in patients with persistent COVID-19 symptoms (10,37). Consistent with findings from our study, previous data has shown that COVID-19 vaccination decreases the risk of PASC and persistent COVID-19 sequelae (23,38); the mechanism underlying this association is unclear. A previous immune phenotyping study of PASC identified increased levels of anti-N and anti-S IgG in individuals with PASC compared to vaccinated controls, indicating persistent chronic immune responses to SARS-CoV-2 viral antigens in PASC, which may be modulated in response to COVID-19 vaccination (34). In agreement with our findings for severe PASC phenotypes, longitudinal evaluations have shown that increasing number of COVID-19 vaccine doses are associated with symptom improvement over time (1,39); however, some studies have suggested that a subset of participants may not experience improvement or may even experience worsening of symptoms after vaccination (40). Further evidence is required to investigate the impact of COVID-19 vaccinations after reinfections, which have been shown to be reduced themselves by booster vaccination (15).

Our study had several strengths including a nationally representative sample of individuals with antibody measurement to detect SARS-CoV-2 N-protein seropositivity, previously diagnosed COVID-19 and reports of long-term sequelae. Furthermore, we were able to assess risk factors related to state-level sociodemographic inequalities, vaccination, reinfection, and predominant variant at the time of infection to explore correlates of PASC in a representative sample of Mexican adults. However, some limitations must be acknowledged to adequately interpret our results. Since the definition of PASC is still evolving, we applied two main definitions based on the World Health organization consensus and a recent definition based on a symptom score (3,4). Despite the robustness of incorporating several definitions of PASC, we were unable to include all symptoms evaluated in the PASC score and symptoms and post-COVID-19 conditions reported in other studies including palpitations, chronic thirst, decreased sexual desire or capacity, abnormal movements, new-onset diabetes, and cardiovascular events including coagulopathy and thrombosis, some of which have previously been reported for cases recovered from severe COVID-19 in Mexico (10); this may lead to an underestimation in the prevalence of severe PASC using this measure and its impact on patients’ health. Next, we used SARS-CoV-2 N-protein seropositivity as a proxy of previous SARS-CoV-2 infection given the high rates of undetected asymptomatic SARS-CoV-2 infections in Mexico, which may otherwise lead to an underestimation of PASC prevalence (41,42). However, a subset of participants who received the CoronaVac inactivated virus vaccine may display seropositivity despite not having been previously infected, this represents 9.44% (95%CI 10.63-11.97) of the total SARS-CoV-2 N-protein seropositive population (43,44). To address this, we also conducted an analysis excluding individuals who received with CoronaVac and individuals who had COVID-19 previously diagnosed by a physician, obtaining largely similar results with slightly lower prevalence estimates (**Supplementary Material**). Finally, although the observed associations are consistent with data from previous studies, its observational nature precludes from making temporal associations with the incidence of PASC, which require further studies to further support these findings in Mexican population.

## Conclusion

We report a prevalence of PASC of 4.67% in the general population, affecting 21.21% of individuals with previously diagnosed COVID-19. In general, over 12.44% of Mexican adults report at least one persistent COVID-19 symptom, with the most common being musculoskeletal pain, headache, cough, loss of smell or taste, and fever. PASC prevalence increased with age and was more common in females; severe PASC was less frequent for cases infected during periods of Omicron variant predominance and in vaccinated individuals in a dose-dependent manner but was more common in individuals with at least one detected SARS-CoV-2 reinfection. Moreover, prevalence of PASC increased in states with higher marginalization, particularly in urban settings. Of note, over 14.05% of participants affected by PASC reported trouble in everyday physical functioning, highlighting the disabling nature of persistent symptoms and indicating a need to implement multidisciplinary teams for patient treatment in Mexico. Further studies are required to characterize the epidemiology of PASC in Mexico using standardized definitions and with longer follow-up in order to identify strategies to mitigate its long-term impact in physical functioning and quality of life as SARS-CoV-2 transitions into endemicity.

## Data Availability

All code, datasets and materials are available for reproducibility of results at http://github.com/oyaxbell/pasc_ensanut/. ENSANUT 2022 data is openly available at: https://ensanut.insp.mx/encuestas/ensanutcontinua2022/descargas.php

https://ensanut.insp.mx/encuestas/ensanutcontinua2022/descargas.php

## ACKNOWLEDGMENTS

CAFM is enrolled at the PECEM Program of the Faculty of Medicine at UNAM. CAFM and DRG are supported by CONACyT.

## AUTHOR CONTRIBUTIONS

Research idea and study design: OYBC, CAFM; data acquisition and processing: CAFM, NEAV, OYBC; statistical analysis: OYBC; analysis/interpretation: CDPC, CAFM, OYBC, NEAV, AVV, LFC, DRG; manuscript drafting: OYBC, CAFM, DRG, LFC, AVV, MRBA, PSC, ANL, NEAV; supervision or mentorship: OYBC, NEAV. Each author contributed important intellectual content during manuscript drafting or revision and accepts accountability for the overall work by ensuring that questions pertaining to the accuracy or integrity of any portion of the work are appropriately investigated and resolved.

## CONFLICT OF INTEREST/FINANCIAL DISCLOSURE

Nothing to disclose.

## FUNDING

This research was supported by Instituto Nacional de Geriatría in Mexico.

## Notes

**CONFLICT OF INTERESTS:** The authors declare that they have no conflict of interests.

### Competing Interest Statement

The authors have declared no competing interest.

### Funding Statement

FUNDING: This research was supported by Instituto Nacional de Geriatria in Mexico.

### Author Declarations

ENSANUT 2022 data is openly available at: https://ensanut.insp.mx/encuestas/ensanutcontinua2022/descargas.php

