## Supplementary Material for "Nationally representative prevalence and determinants of post-acute sequelae of SARS-CoV-2 infection (Long COVID) amongst Mexican adults in 2022"

**SUPPLEMENTARY METHODS**

**National Health and Nutrition Survey**

The Mexican National Health and Nutrition Survey (ENSANUT) comprises a series of population-based surveys that provide precise information on the health and nutritional status of Mexican population, as well as on the performance of our health care system. ENSANUT uses a two-stage probabilistic cluster stratified sampling based on households and individuals from each group of interest (adults [≥20 years], adolescents [10-19 years], school age [5-9 years], and preschool age [0-4 years]), and it is representative at a regional (North, Center, Mexico City, and South), and national level for rural and urban areas. ENSANUT has been conducted in 2006, 2012, 2016, 2018, 2020, 2021, and 2022. Sociodemographic and dwelling data is obtained from each household, and individual participants undergo a standardized health questionnaire that is applied in face-to-face interviews, followed by a physical exam including measurement of blood pressure and anthropometry (height in centimeters, weight in kilograms, and waist circumference in centimeters [not available for 2020]) by trained personnel. The physical exam was performed only in a subset of participants for the 2018 (40%) and 2021 (80 %) surveys. Additionally, a random subset of each cycle was selected for venous blood collection for biochemical evaluation of the following serum laboratory parameters: albumin, creatinine, uric acid, triglycerides, total cholesterol, LDL-cholesterol, HDL-cholesterol, glucose, insulin, glycated hemoglobin, and from 2020 onwards SARS-CoV-2 serology. All questionnaires and procedures were reviewed and approved by the Ethics, Investigation and Biosecurity Committee from the National Institute of Public Health for each year of survey (1–4).

**SUPPLEMENTARY TABLES**

**SUPPLEMENTARY TABLE 1.** Sociodemographic and clinical characteristics of individuals with post-acute sequelae of SARS-CoV-2 symptoms (PASC) as identified by a PASC score ≥12 compared with PASC indeterminate individuals in ENSANUT 2022.

|  | Overall Sample | | |  |
| --- | --- | --- | --- | --- |
| Characteristic | Overall, N = 5,211^1^ | PACS indeterminate, N = 4,766^1^ | PACS positive, N = 445^1^ | p-value^2^ |
| **Male sex (%)** | 2,322 (45%) | 2,124 (45%) | 198 (44%) | >0.9 |
| **Age (years)** | 43 (31, 55) | 43 (31, 55) | 47 (34, 56) | 0.007 |
| **Reinfection (%)** | 638 (12%) | 584 (12%) | 54 (12%) | >0.9 |
| **Omicron variant (%)** | 2,087 (40%) | 1,954 (41%) | 133 (30%) | <0.001 |
| **COVID-19 vaccine (%)** | 4,830 (93%) | 4,442 (93%) | 388 (87%) | <0.001 |
| **Symptom duration** |  |  |  | <0.001 |
| <1 month | 1,363 (47%) | 1,113 (46%) | 250 (56%) |  |
| 1-3 months | 478 (17%) | 382 (16%) | 96 (22%) |  |
| 3-6 months | 124 (4.3%) | 107 (4.4%) | 17 (3.8%) |  |
| >6 months | 126 (4.4%) | 109 (4.5%) | 17 (3.8%) |  |
| Still persistent | 800 (28%) | 735 (30%) | 65 (15%) |  |
| **Any symptom (%)** | 2,839 (54%) | 2,394 (50%) | 445 (100%) | <0.001 |
| **Loss of smell/taste (%)** | 692 (13%) | 336 (7.0%) | 356 (80%) | <0.001 |
| **Brain fog (%)** | 166 (3.2%) | 68 (1.4%) | 98 (22%) | <0.001 |
| **Gastrointestinal symptoms (%)** | 160 (3.1%) | 77 (1.6%) | 83 (19%) | <0.001 |
| **Chest pain (%)** | 285 (5.5%) | 146 (3.1%) | 139 (31%) | <0.001 |
| **Breathlessness (%)** | 358 (6.9%) | 221 (4.6%) | 137 (31%) | <0.001 |
| **Dyspnea (%)** | 513 (9.8%) | 349 (7.3%) | 164 (37%) | <0.001 |
| **Musculoskeletal pain (%)** | 913 (18%) | 645 (14%) | 268 (60%) | <0.001 |
| **Dizziness (%)** | 213 (4.1%) | 108 (2.3%) | 105 (24%) | <0.001 |
| **Headache (%)** | 817 (16%) | 586 (12%) | 231 (52%) | <0.001 |
| **Weight loss (%)** | 203 (3.9%) | 104 (2.2%) | 99 (22%) | <0.001 |
| **Loss of appetite (%)** | 210 (4.0%) | 106 (2.2%) | 104 (23%) | <0.001 |
| **Kidney problems (%)** | 55 (1.1%) | 22 (0.5%) | 33 (7.4%) | <0.001 |
| **Sleep disturbances (%)** | 288 (5.5%) | 166 (3.5%) | 122 (27%) | <0.001 |
| **Fever (%)** | 670 (13%) | 446 (9.4%) | 224 (50%) | <0.001 |
| **Depression (%)** | 187 (3.6%) | 107 (2.2%) | 80 (18%) | <0.001 |
| **Anxiety (%)** | 256 (4.9%) | 153 (3.2%) | 103 (23%) | <0.001 |
| **Fatigue (%)** | 1,250 (24%) | 952 (20%) | 298 (67%) | <0.001 |
| **Chronic cough (%)** | 978 (19%) | 645 (14%) | 333 (75%) | <0.001 |
| **Post-exertional malaise (%)** | 541 (10%) | 268 (5.6%) | 273 (61%) | <0.001 |
| **PASC score** | 0.0 (0.0, 5.0) | 0.0 (0.0, 4.0) | 15.0 (13.0, 19.0) | <0.001 |
| **DISLI quartile** |  |  |  | 0.003 |
| Q1 | 1,707 (33%) | 1,594 (33%) | 113 (25%) |  |
| Q2 | 1,630 (31%) | 1,465 (31%) | 165 (37%) |  |
| Q3 | 965 (19%) | 882 (19%) | 83 (19%) |  |
| Q4 | 909 (17%) | 825 (17%) | 84 (19%) |  |
| **CES-D score** | 5.0 (3.0, 8.0) | 5.0 (3.0, 8.0) | 6.0 (3.0, 9.0) | 0.075 |
| **Diabetes (%)** | 358 (14%) | 311 (14%) | 47 (21%) | 0.002 |
| **Hypertension (%)** | 505 (20%) | 447 (20%) | 58 (26%) | 0.018 |
| **Daily smoking (%)** | 178 (7.1%) | 164 (7.2%) | 14 (6.3%) | 0.6 |
| **Vaccine type** |  |  |  | <0.001 |
| Unvaccinated | 381 (7.3%) | 324 (6.8%) | 57 (13%) |  |
| mRNA | 1,607 (31%) | 1,476 (31%) | 131 (29%) |  |
| Adenovirus vector | 2,629 (50%) | 2,422 (51%) | 207 (47%) |  |
| Inactivated virus | 438 (8.4%) | 397 (8.3%) | 41 (9.2%) |  |
| Other | 156 (3.0%) | 147 (3.1%) | 9 (2.0%) |  |
| **Incapacitating symptoms** | 457 (8.8%) | 245 (5.1%) | 212 (48%) | <0.001 |
| ^1^n (%); Median (IQR) | | | | |
| ^2^Pearson's Chi-squared test; Wilcoxon rank sum test; Fisher's exact test | | | | |

**SUPPLEMENTARY FIGURES**

**Supplementary Figure 1.** Flowchart of participant selection for ENSANUT 2022. We display the total number of participants surveyed per year (first row). The ***general population*** of our study is therefore comprised of participants ≥20 years old that completed the health questionnaire (second row), while the ***laboratory subset*** includes those with complete weights for venous blood and sampling stratum (third row). A subset of analyses was conducted in individuals with previously diagnosed COVID-19 and complete data (fourth row). This diagram was designed using resources created by *Freepik* from [www.flaticon.com](http://www.flaticon.com)

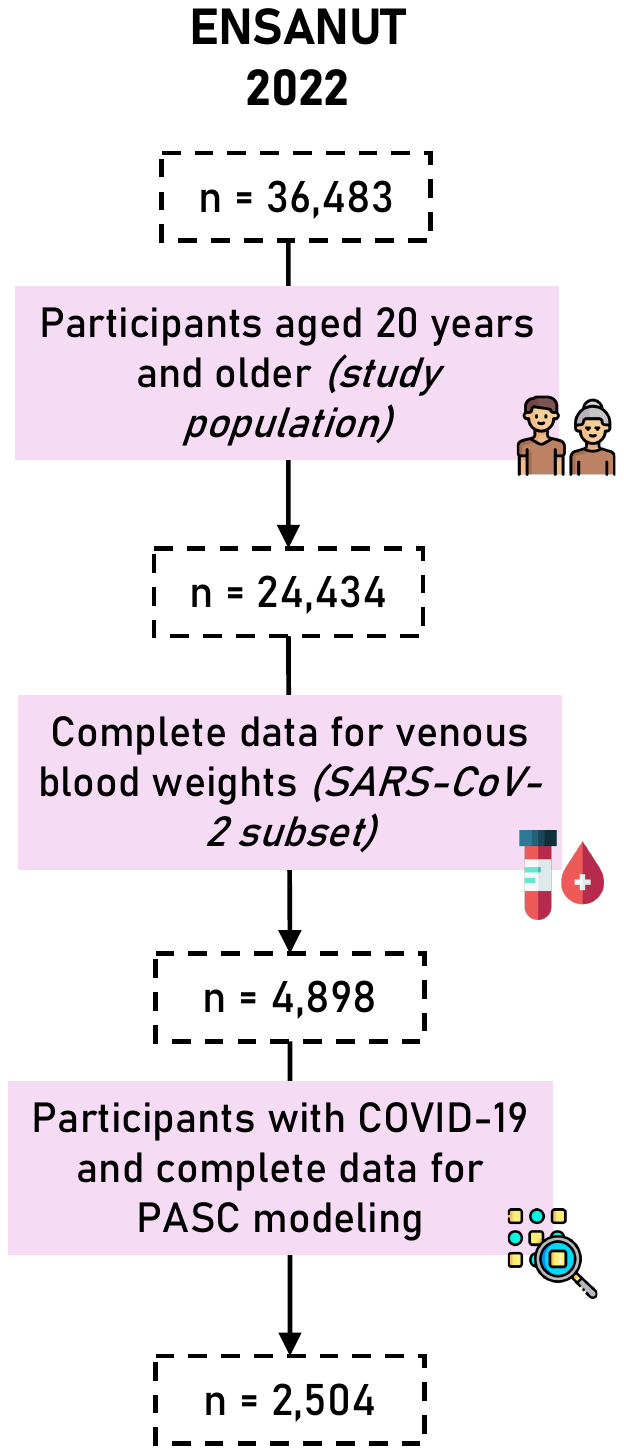

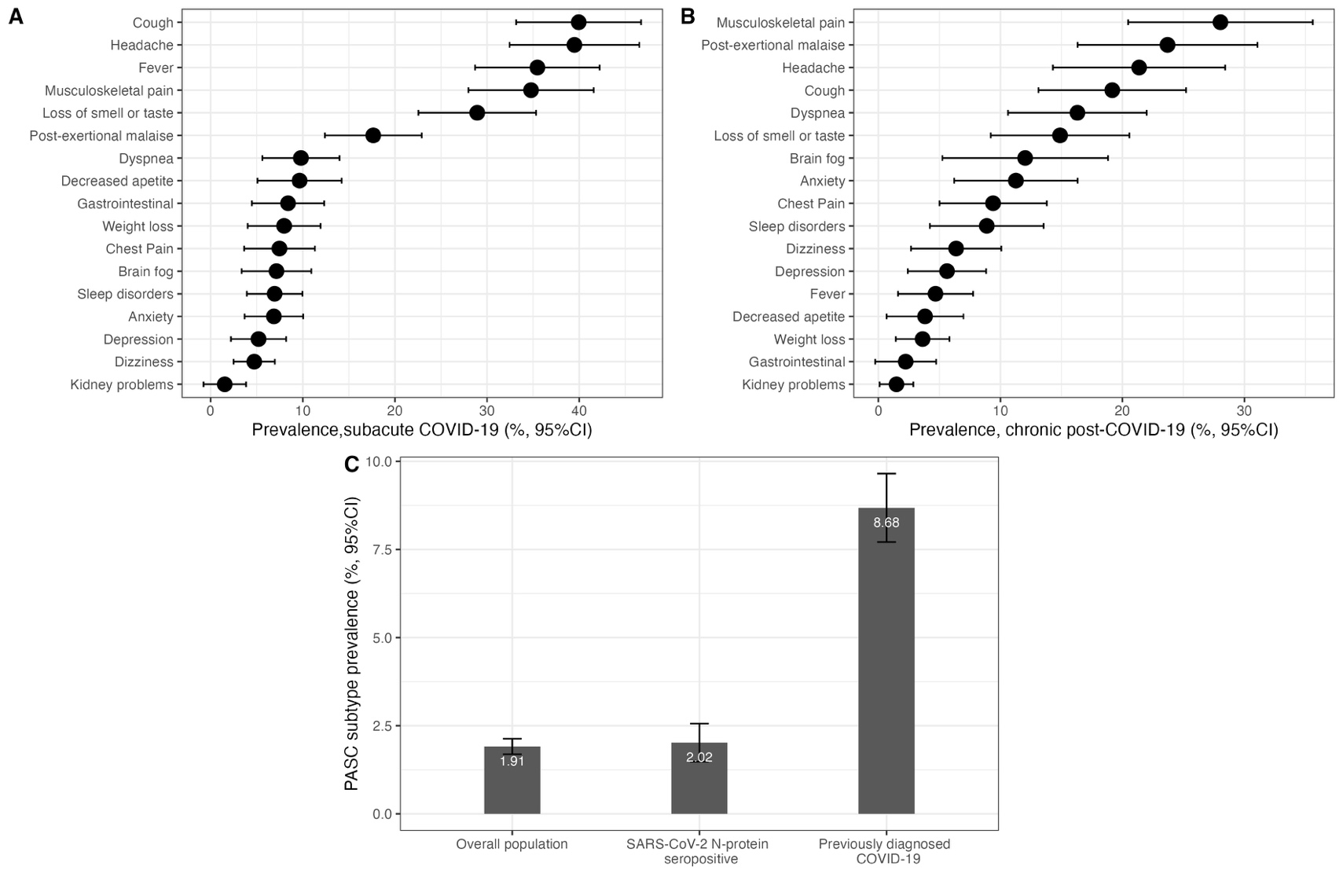

**Supplementary Figure 2.** Prevalence of persistent COVID-19 symptoms amongst SARS-CoV-2 N-protein seropositive adults ≥20 years in ENSANUT 2022 stratified comparing cases with subacute or ongoing symptomatic COVID-19 (**A**) and chronic post-COVID-19 syndrome (**B**). The figure also shows the prevalence of PASC using the definition of PASC score ≥12 points amongst the overall, seropositive and previously diagnosed with COVID-19 population (**C**).

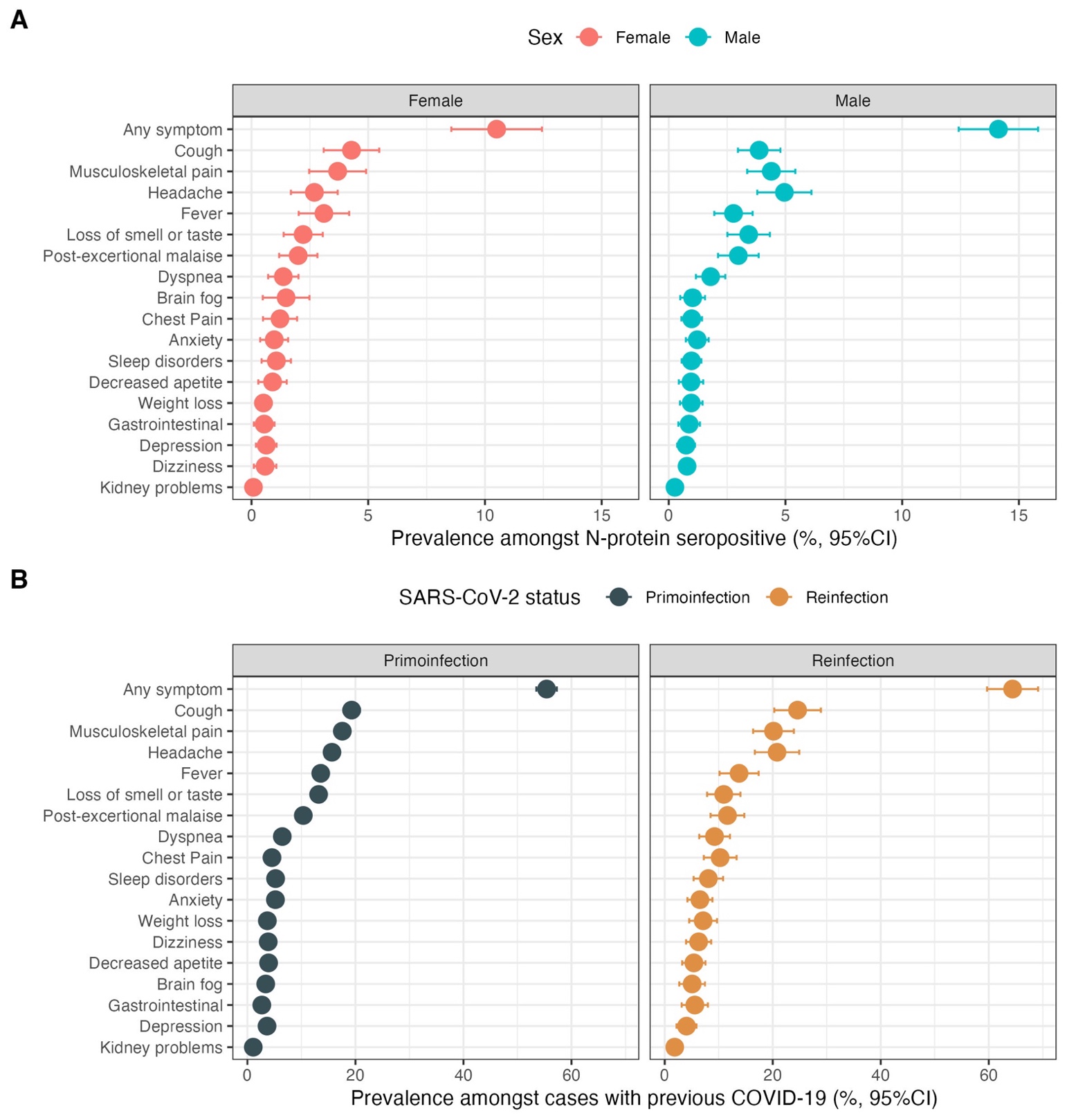

**Supplementary Figure 3.** Prevalence of persistent COVID-19 symptoms amongst SARS-CoV-2 N-protein seropositive adults ≥20 years in ENSANUT 2022 stratified by sex (A) and by SARS-CoV-2 infection status (primoinfection vs. at least one reinfection).

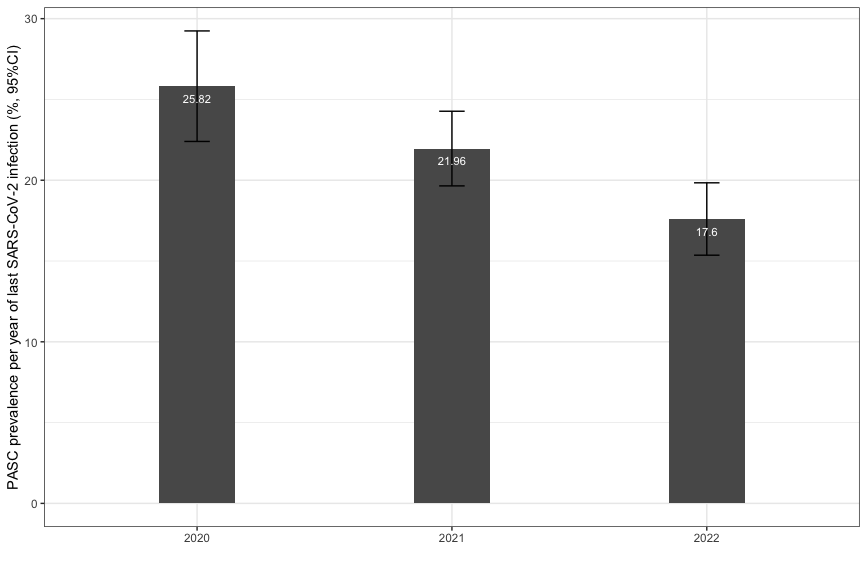

**Supplementary Figure 4.** Comparison of the prevalence of post-acute sequelae after SARS-CoV-2 infection (PASC) for cases with infections detected in 2020, 2021 and 2022.

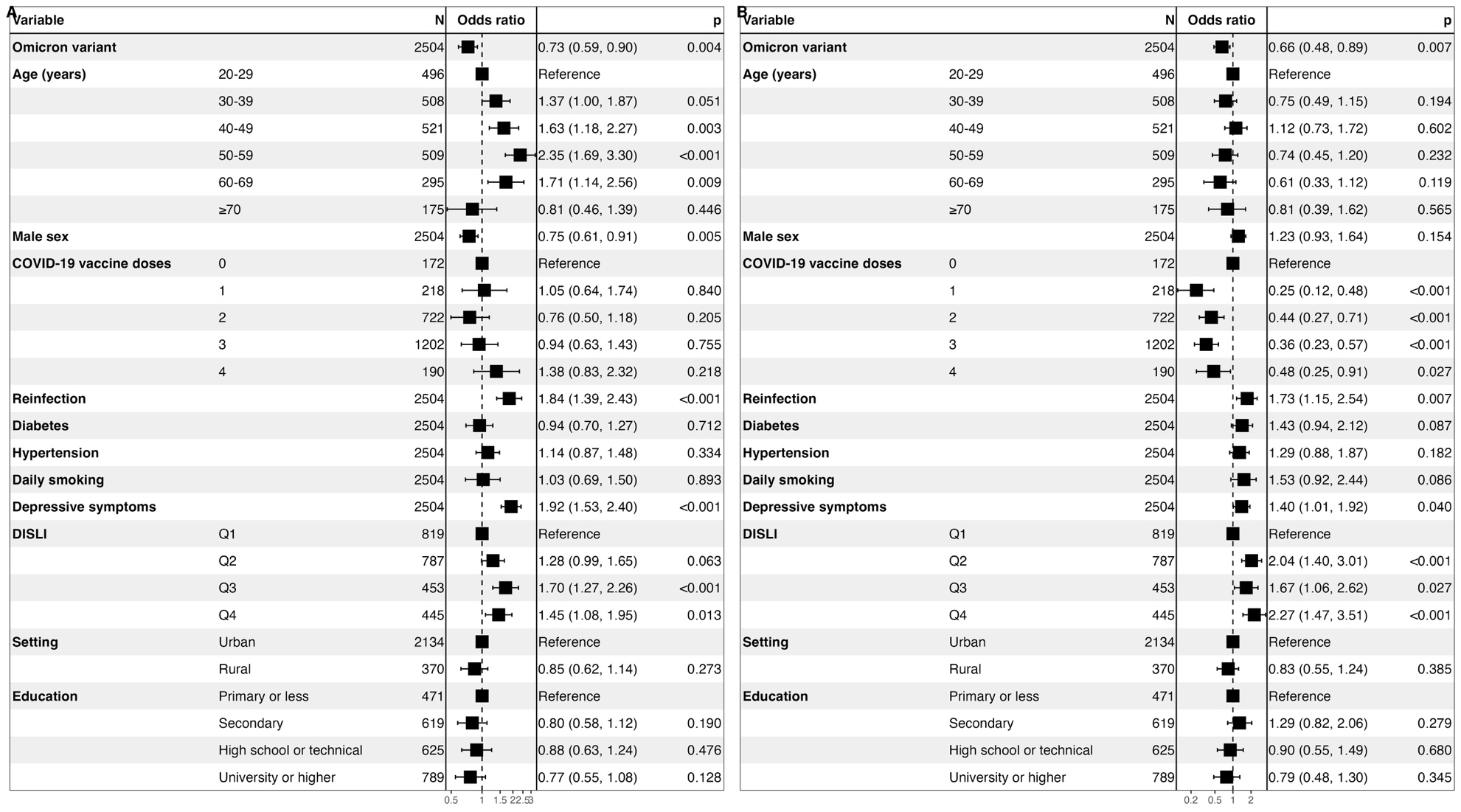

**Supplementary Figure 5.** Fixed effects logistic regression model adjusted by survey weights for prediction of post-acute sequelae of SARS-CoV-2 symptoms (PASC) using the WHO definition (**A**) and identified by a PASC score ≥12 (**B**) amongst Mexican adults enrolled in ENSANUT 2022, disaggregating vaccinations by the number of vaccine doses compared to unvaccinated individuals.

**Abbreviations:** DISLI, Density-independent social lag-index; Depressive symptoms: Moderate to severe depressive symptoms identified by the Center for Epidemiologic Studies Depression Scale

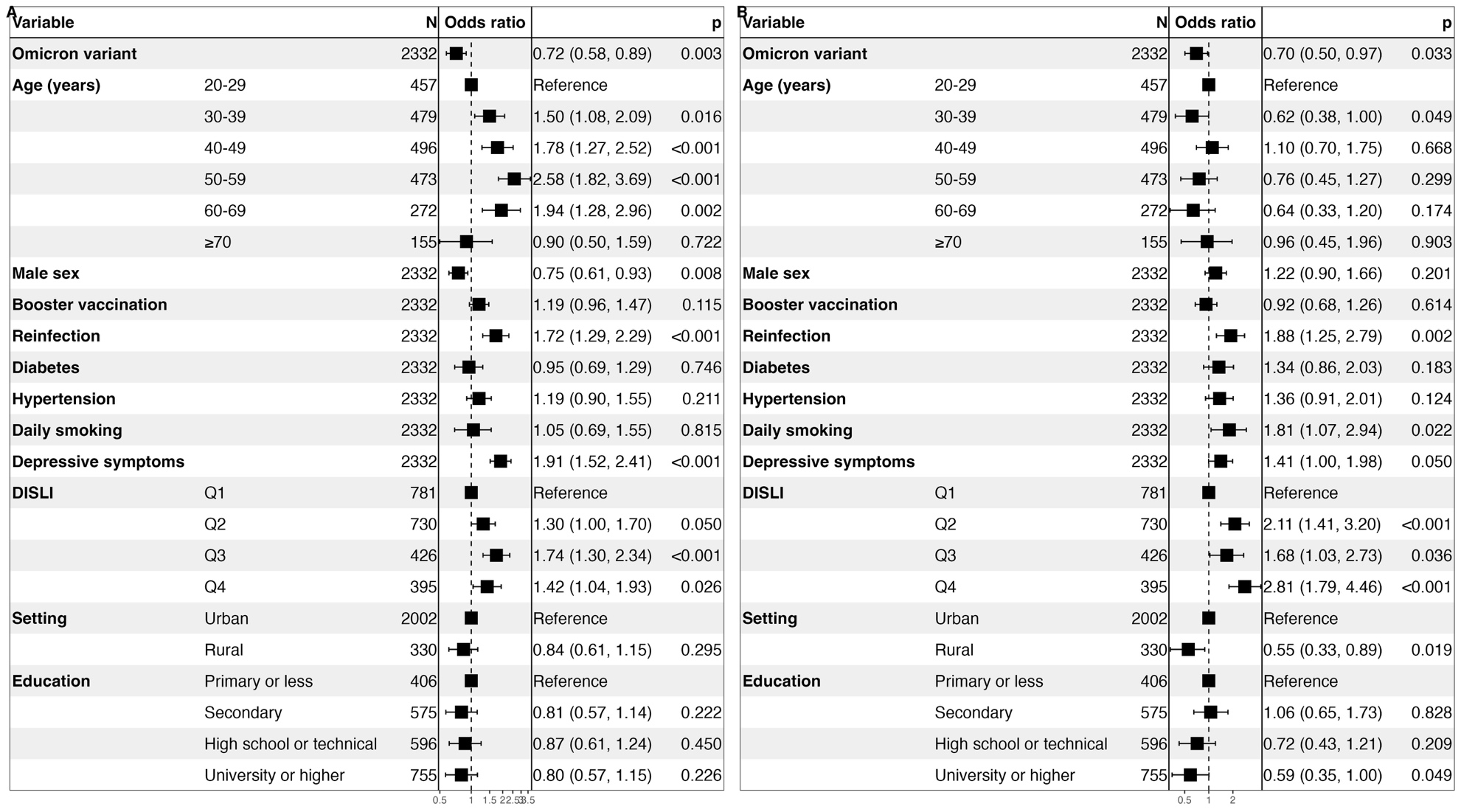

**Supplementary Figure 6.** Fixed effects logistic regression model adjusted by survey weights for prediction of post-acute sequelae of SARS-CoV-2 symptoms (PASC) using the WHO definition (**A**) and identified by a PASC score ≥12 (**B**) amongst Mexican adults enrolled in ENSANUT 2022 who received full primary vaccination schedule to assess the influence of vaccine boosters.

**Abbreviations:** DISLI, Density-independent social lag-index; Depressive symptoms: Moderate to severe depressive symptoms identified by the Center for Epidemiologic Studies Depression Scale

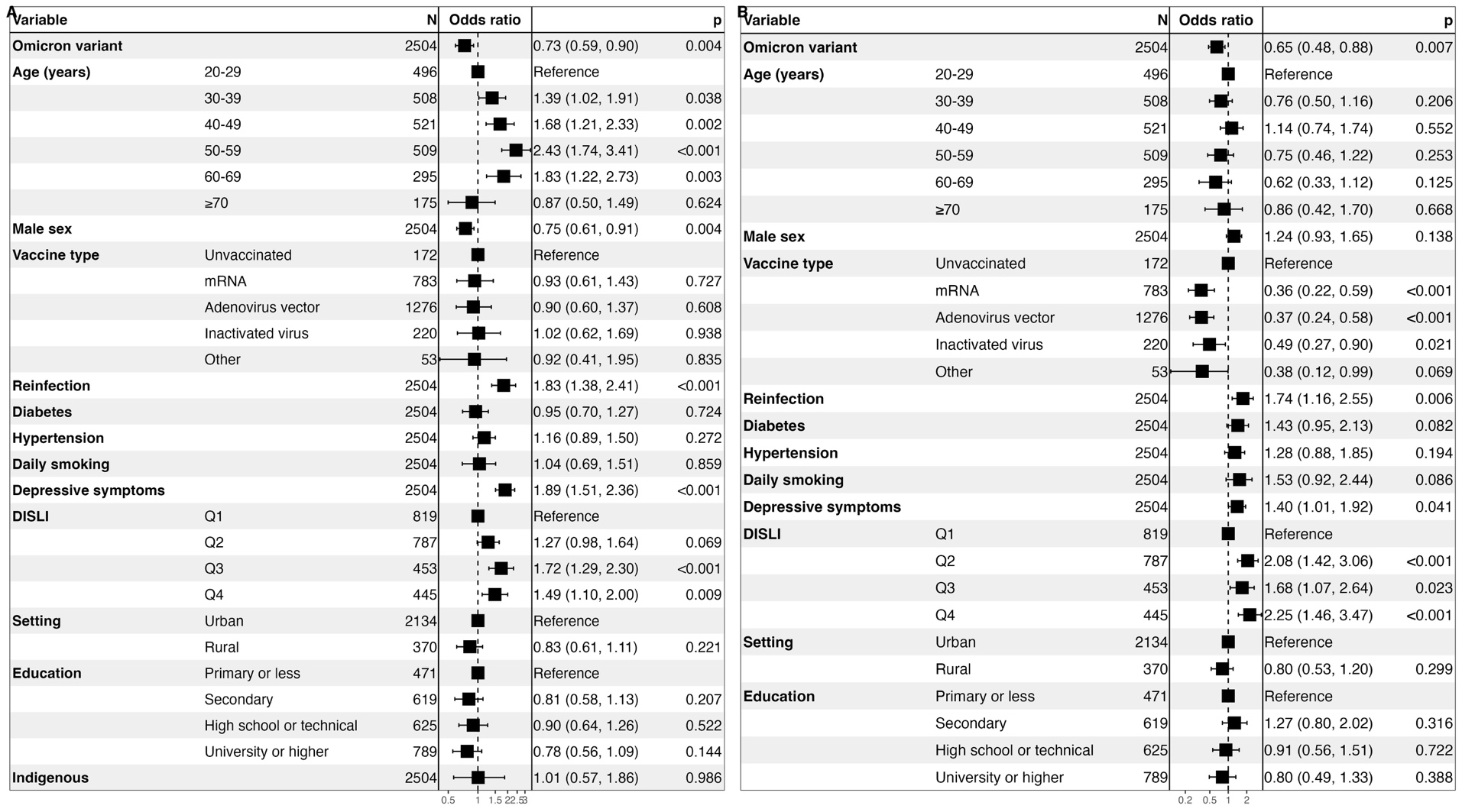

**Supplementary Figure 7.** Fixed effects logistic regression model adjusted by survey weights for prediction of post-acute sequelae of SARS-CoV-2 symptoms (PASC) using the WHO definition (**A**) and identified by a PASC score ≥12 (**B**) amongst Mexican adults enrolled in ENSANUT 2022, disaggregating vaccinations by vaccine type of primary vaccination schedule compared to unvaccinated individuals.

**Abbreviations:** DISLI, Density-independent social lag-index; Depressive symptoms: Moderate to severe depressive symptoms identified by the Center for Epidemiologic Studies Depression Scale

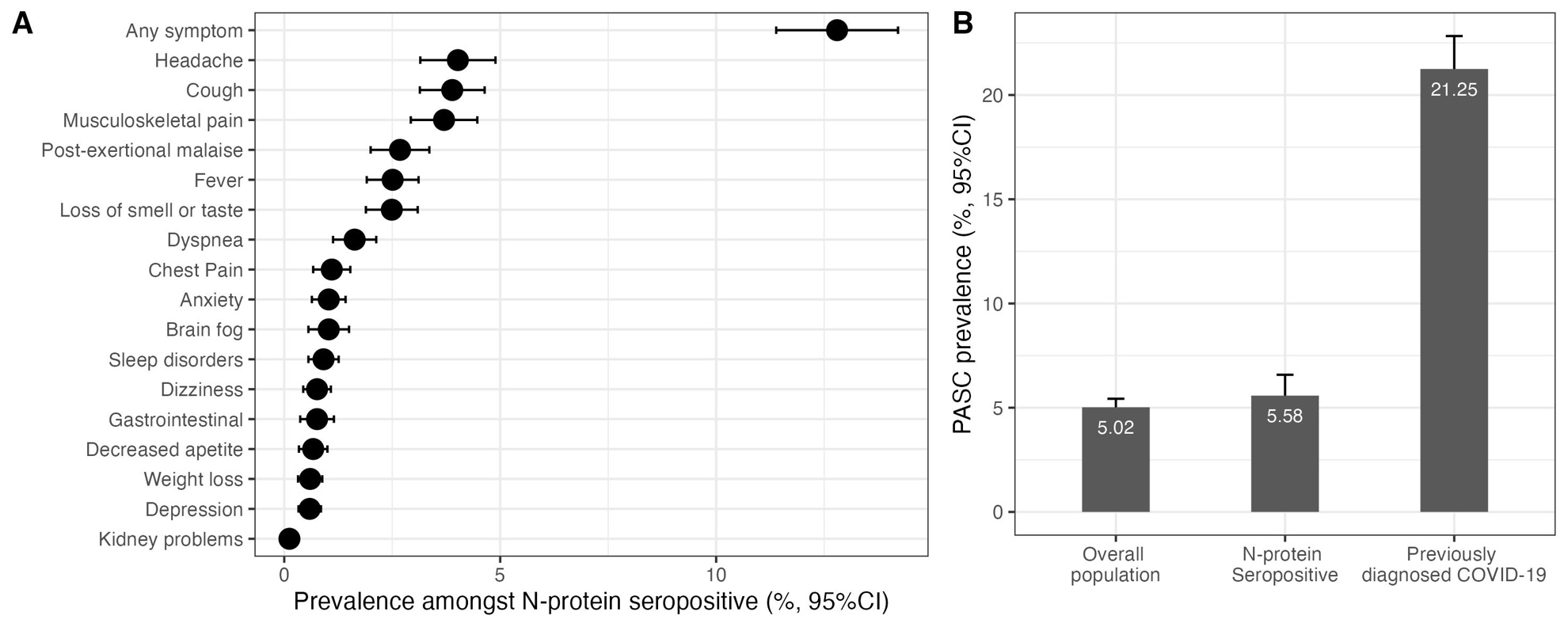

**Supplementary Figure 8**. Prevalence of persistent COVID-19 symptoms amongst SARS-CoV-2 N-protein seropositive adults ≥20 years in ENSANUT 2022 who did not receive the CoronaVac vaccine (A) and prevalence of post-acute sequelae of SARS-CoV-2 symptoms (PASC) as identified by the WHO definition in the overall population who did not receive the CoronaVac vaccine, in SARS-CoV-2 N-protein seropositive adults who did not receive the CoronaVac vaccine and in individuals with COVID-19 previously diagnosed by a physician who did not receive the CoronaVac vaccine in the ENSANUT 2022 sample (B).
